# Transdiagnostic neurocognitive endophenotypes in major psychiatric illness

**DOI:** 10.1101/2020.02.14.20022863

**Authors:** Bharath Holla, Pavithra Dayal, Aswathy Das, Mahashweta Bhattacharya, V Manjula, Dhruva Ithal, Srinivas Balachander, Jayant Mahadevan, Ravi Kumar Nadella, Vanteemar S Sreeraj, Vivek Benegal, Janardhan Y. C. Reddy, Urvakhsh Meherwan Mehta, ADBS Consortium, Biju Viswanath

**Author notes:** **Corresponding author** Dr. Biju Viswanath, Associate Professor of Psychiatry, National Institute of Mental Health and Neurosciences, Bangalore, India 560029.

## Abstract

We aimed to characterize potential transdiagnostic neurocognitive endophenotypes in a large cohort of multiplex families affected with two or more individuals having a major psychiatric illness(MPI) i.e., schizophrenia(SCZ), bipolar disorder(BPAD), substance use disorders(SUD) and obsessive-compulsive disorder(OCD). We recruited 640 individuals that included 256 affected individuals with MPI, 227 of their unaffected first-degree relatives(FDR) and 157 population healthy-controls(PHC). Neurocognitive battery included assessments of executive function, working memory, social cognition, verbal learning and recall. Linear mixed effects models were applied to the neurocognitive components to examine their transdiagnostic and endophenotype status after accounting for demographic and family variables. We also examined the relationship of cognitive domains with diagnosis-specific Family History Density score(FHD).

The deficits in cognitive flexibility, working memory and social cognition were transdiagnostic; processing speed was impaired in SCZ and BPAD groups while verbal learning and recall in SCZ, BPAD and SUD groups. These deficits with the exception of social cognition, worsened with age and parental education had protective effect. The unaffected FDRs had deficits in all the domains except processing speed in comparison to PHC; social cognition deficits were comparable to affected individuals. The diagnosis-specific FHD analysis showed that deficits in cognitive flexibility, working memory and social cognition were endophenotypes across disorders.

Evaluation of neurocognitive functions across multiple affected individuals in a large multiplex family-based cross-disorder cohort, has the potential to elucidate transdiagnostic as well as endophenotype vulnerabilities to psychiatric illness. This study adds to the emerging conceptualization of psychiatric illness as a combination of both diagnosis-specific and transdiagnostic markers.

## Introduction

Major psychiatric illnesses are highly heritable, yet the search for replicable genetic and neurobiological mechanisms underlying them has proven highly challenging by [1]. One of the major reasons for this non-replicability is the focus on discrete clinical disease constructs, as categorized in current classificatory systems (ICD-10 & DSM5) by their “behavioral exophenotype” or phenomenology. There exists wide within-diagnoses heterogeneity in clinical phenomenology, as well as, frequent comorbidities [2]. The specificity of familial transmission for several psychiatric diagnosis is only partial [3,4]. More importantly, extant research has shown that there is considerable overlap in the genetic and endophenotypic markers of these disorders [5], pointing towards a common “transdiagnostic mechanism” for vulnerability to mental illnesses, which works in combination with diagnosis specific mechanisms. Large scale genomic studies have identified significant overlaps in the genetic risks, cutting across traditional diagnostic boundaries [6,7]. Transdiagnostic neuroanatomical endophenotypes have also been examined in one recent study [8].

Neurocognitive functions, including the general cognitive abilities (or ‘g’) are highly heritable [9] and have been studied extensively as endophenotypic markers for psychiatric illnesses [10,11]. This approach has also led to discovery of several promising candidate genes of influence [12]. Impairments in working memory, executive functions, set-shifting, decision making, response inhibition and social cognition have been consistently found in affected probands and to varying degree in their unaffected first-degree relatives (FDR) [13–23]. However, nearly all extant research in this area has included individuals from non-multiplex families, and looked at endophenotypes only when the probands are affected with a single, distinct psychiatric illness. The use of individualized neuropsychological batteries for various disorders by different study groups, makes it difficult to examine which of these deficits are shared between psychiatric disorders, and which of them are distinct.

Studying large numbers of multiplex families with affected members having several psychiatric illnesses, and unaffected FDRs, would provide the most statistically robust method for the identification of neurocognitive endophenotypes that cut across traditional diagnostic boundaries[5]. Specifically, research in multiplex families would reduce the potential of spurious or non-genetic forms of the illness, and increase the potential that endophenotypes identified reflect genetic factors that influence disease vulnerability [24]. This approach has been adopted in our ongoing longitudinal study, the Accelerator Program for Discovery in Brain Using Stem Cells (ADBS), in which we recruit families in whom multiple members (≥ 2 affected FDRs in a nuclear family) are diagnosed to have a major psychiatric disorder [schizophrenia (SCZ), bipolar disorder (BPAD), obsessive compulsive disorder (OCD), Alzheimer’s dementia and substance use disorder (SUD)] [4][25]. We report here the neurocognitive functioning in affected (N=256) and unaffected (N=227) FDRs in these multiplex families in comparison with population healthy controls (PHC) (N=157), who do not have family history of major psychiatric illness. Patients with dementia of Alzheimer’s type (DAT) were excluded from this analysis as cognitive deficits are defining features of dementias. We hypothesized that a common transdiagnostic neurocognitive deficit can be identified in major psychiatric illnesses, in addition to disorder specific deficits. We also hypothesized that these neurocognitive dysfunctions would, at least in part, be evident in unaffected FDRs and hence reflect pre-existing familial endophenotypic vulnerability. We expected some of these deficits to be transdiagnostic endophenotypes, and others to be predicted by diagnosis specific family history density (FHD). Finally, we also hypothesized that these deficits would worsen with greater disease severity.

## METHODS

### Subjects

The study sample included patients diagnosed with four major psychiatric illnesses (MPI) according to the DSM-IV criteria namely SCZ (N=68), BPAD (N=65), OCD (N=66) and SUD (N=57), their unaffected FDR (n= 227), and population healthy controls (PHC) (n= 157). All affected probands were recruited from the adult psychiatry services and specialty psychiatry clinics of NIMHANS, Bengaluru. The healthy comparison group (PHC) were recruited from the community by word-of-mouth, local advertisements and from a previous study conducted at the same centre with similar methodology [26]. The recruited patients had a strong family history with at least another affected FDR with one of the 4 MPIs, as ascertained by Family Interview for Genetic Studies (FIGS)(Figure 1A)[27]. The subjects in the FDR group did not have any of the four listed diagnoses (n=197), but may have had a life-time diagnosis of other psychiatric diagnosis as per MINI, such as dysthymia or other anxiety spectrum disorders (n=30) (Figure 1B). The PHC did not have any Axis 1 psychiatric diagnosis (as per MINI) and had no family history of MPI in their FDR. All the DSM-IV psychiatric diagnoses were corroborated by two trained psychiatrists using the Mini International Neuropsychiatric Interview (MINI) [28]. The study protocol and recruitment procedures are detailed elsewhere [4,25]. The NIMHANS ethics committee approved the study protocol and written informed consent was obtained from all the participants in the study. (Demographic and clinical characteristics are shown in Supplementary Material Table 1)

**Figure 1:**
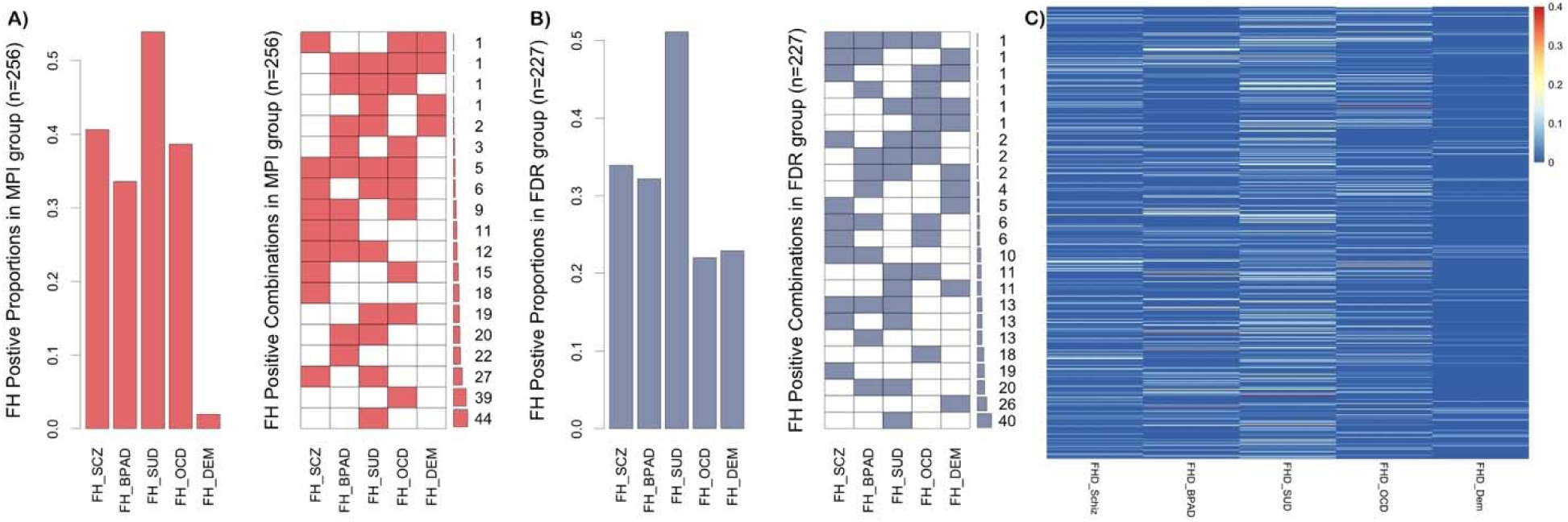
Representation of presence of family history in the sample. A) Proportion and Combination of Family History in the Major Psychiatric Illness group. B) Proportion and Combination of Family History in First-Degree Relatives. C) Heat plot of Family History Density Score. FH, Family History; MPI, Major Psychiatric Illness; FDR, First Degree Relatives; SCZ, schizophrenia; BPAD, bipolar affective disorder; OCD, obsessive compulsive disorder; SUD, substance use disorder; DEM, dementia.

### Neuropsychological Assessments

All neurocognitive tests were administered to participants by trained psychologists, in a quiet room free of distraction, during a single session that lasted approximately 40 to 50 minutes on the forenoon of the assessment day. The order of the administration was counter balanced. The following 4 tests were chosen based on the existing literature indicating varying levels of deficits in these domains in the four disorders under study and importantly, these tests were less sensitive to the influences of language and cultural bias. **Color Trails (CT)Tests** [29] A and B (an analogue of the Trail Making Test), was used to evaluate speed of processing and cognitive flexibility. Total time taken to complete the test A and B were recorded in seconds along with errors calculated from the misses and wrong selection. An “effective time-per-circle” measure was calculated using the formula [TT/CC+TE/TC], where TT is Total time taken, CC is number of circles completed, TE is Total Errors committed, and TC is total circles in the test (ie., 25 for CT-A and 50 for CT-B). One-back and 2 back versions of the **Auditory N-back** test were used to assess verbal working memory [30]. Thirty randomly ordered consonants which are common in Indian languages were presented [31]. Total number of hits and errors (omissions and commissions) were recorded. Based on signal detection theory, a sensitivity index (d’) was calculated for each of 1- and 2-back tests, as a difference between the Z transforms of Hit Rates (HR) and False Alarm rates (FA), d’ = z(HR) – z(FA) [32], where Hit Rate is Hits/Targets and FA is Commission Errors/Distractors. Perfect hits (ie., HR=1) were adjusted using the formula [1 − 1/(2n)], where n is total number of targets (n=9) and zero FA scores were adjusted using the formula [1/(2n)], where n is total number of distractors (n=21). **Rey’s Auditory Verbal Learning Test (RAVLT)** was used to assess verbal learning and recall [33]. It consists of two lists A and B with 15 different words from familiar objects like vehicles, tools, animals and body parts. The words were translated in to Indian languages (Hindi, Kannada, Telugu, Tamil and Malayalam)[31]. Total numbers of words correctly recalled over all the five trials (learning score) and the number of words recalled correctly in the immediate recall trial were recorded. Since the delayed-recall scores correlate highly with total scores (r >.75) [34] these were not recorded, in the interest of time. **Social Cognition Rating Tools in Indian Setting (SOCRATIS)** was used to assess social cognition in the Indian socio-cultural setting [35]. Subtests from SOCRATIS were used to assess second order theory of mind, since this measure showed consistent associations with real-world outcomes [36,37], as well as, neural signatures [26] in earlier studies. Adapted versions of two false belief stories (Ice-cream man task [38] and hidden banana task [39]) and two Irony detection stories [40] were administered. A second order theory of mind index [35] was calculated as the proportion of correct responses on these tests after accounting for responses to control questions.

The neurocognitive scores were summarized into five domains of i) Processing speed (CTT-A effective time-per-circle), ii) Cognitive flexibility (CTT-B effective time-per-circle), iii) Working memory (auditory sensitivity index for 1-back and 2-back d’ scores), iv) Verbal learning and recall (total learning and immediate recall scores), and v) Social cognition (2^nd^ order theory of mind task).

### Family History Density

As we had multiplex families with at least 2 FDRs affected with one of the severe mental illness, we calculated Family History Density (FHD) which was proposed by Stoltenberg [41] for alcohol use disorder. Weighted points were given to the family members based on their relatedness to the subject (parents (0.5) and grandparents (0.25)) if they are affected and unaffected relatives were given a score of zero. These scores were summed to obtain the FHD score for respective illness (explained in detail elsewhere [4]). This score was later used in regression analysis with the neurocognitive measures.

### Statistical Analysis

Missing data for neurocognitive variables were imputed when no more than 3 of the 7 test variables were missing per subject. Among the 640 subjects, 599 (93.56%) had no missing data (See Supplementary Figure 1 for missingness report). Missing data were imputed using multivariate imputation by chained equations (MICE) with a random forest-based algorithm to produce an unbiased estimate with a narrow confidence interval [42]. The raw scores from the five cognitive domains scores were transformed to T-scores (mean = 50; standard deviation = 10). In order to make the CTT-A and CTT-B scores directionally consistent, they were reversed by multiplying it with −1, thus high values for all cognitive domains indicated good performance. For domains with more than one measure, the individual T scores were summed and then the sum was standardized to a T score.

Separate linear mixed effects models were implemented for each neurocognitive domain with group status as the fixed effect predictor. Additionally, to address the potentially confounding effects of demographic variables, we added age, gender, parental education (as fixed effects predictor) and family ID (as random effects predictor). In order (i) to evaluate if the deficits were transdiagnostic, we compared the component scores for each of the primary diagnoses to the PHC group. If the deficits were noted in all the comparisons, then the component was deemed as transdiagnostic or as diagnosis specific if otherwise, and (ii) to evaluate the endophenotype status of these components, we first compared them across the three groups (PMI, FDR and PHC). A mixed-effect multivariate multiple regression was fit to examine the relationship between specific familial risk and neurocognitive domains. Lastly, we explored the effect of illness severity (illness duration and clinical global impression severity score) on cognitive domains using Pearson correlation. To ensure comparability across different diagnoses, illness duration was normalized within each diagnostic group using the formula: (individual duration□−□minimum duration)/(maximum duration □−□minimum duration). All analyses were performed in R environment for statistical computing, version 3.5.2, using the packages mice [43], psych [44], and lme4 [45].

## RESULTS

### Transdiagnostic evaluation of the Neurocognitive Domains

Linear Mixed Effects Evaluation of the neurocognitive domains (in Supplementary Material Table 2) revealed diagnosis-specific deficits for processing speed with only SCZ and BPAD group showing deficits when compared to healthy controls. In the verbal learning and recall domain, the SCZ, BPAD and SUD groups showed deficits but not the OCD group. In the cognitive flexibility, working memory and social cognition domains the deficits were transdiagnostic, with all groups showing deficits when compared to healthy controls. Figure 2 shows the estimated marginal means ± 95% confidence interval for the neurocognitive domains among different groups.

**Figure 2:**
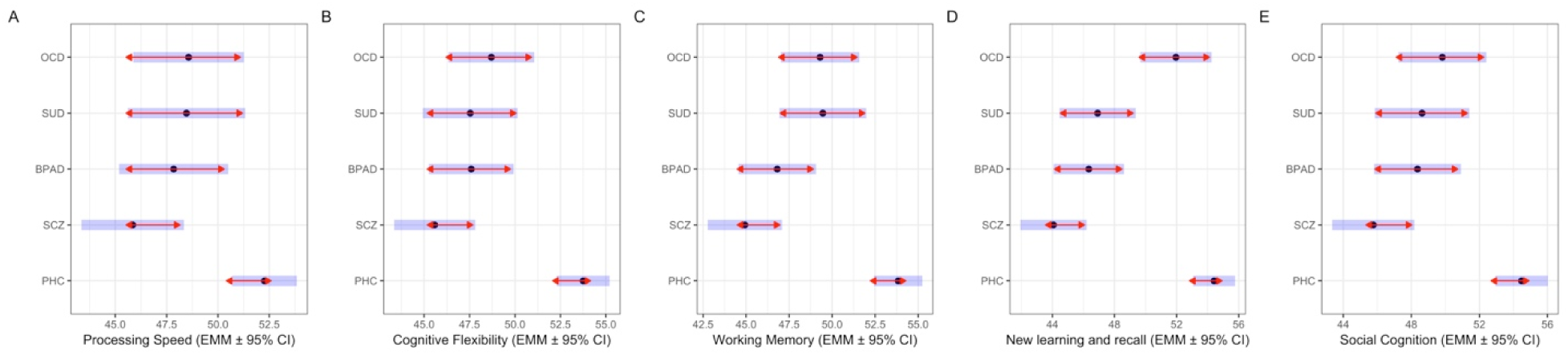
Estimated Marginal Means ± 95% Confidence Interval (CI) for the Neurocognitive domains among Major Psychiatric Illness and Population Healthy Controls. A)Processing speed. B)Cognitive Flexibility. C)Working Memory. D)Verbal(New) Learning and Recall. E)Social cognition PHC, population healthy control; SCZ, schizophrenia; BPAD, bipolar affective disorder; OCD, obsessive compulsive disorder; SUD, substance use disorder; EMM, estimated marginal means; CI, confidence interval.

Further all neurocognitive domains with the exception of social cognition, worsened with age, while parental education showed protective effects. Males performed poorly on verbal learning and recall domain but had comparable performance in all other domains. About 42.3%, 30.1%, 32%, 53.8% and 31.7% variance was explained by the models for processing speed, cognitive flexibility, working memory, verbal learning and recall and social cognition domains respectively as noted by conditional R^2^ (with fixed and random effects predictors) [46].

### Endophenotypic evaluation of the Neurocognitive Domains

To elucidate whether the diagnosis-specific and transdiagnostic deficits in the neurocognitive components were marker of current psychiatric illness or if they were reflective of pre-existing familial endophenotypic vulnerability, we implemented a linear-mixed effects model to compare the component scores among FDR of patients with MPI and PHC, accounting for demographic and family ID as stated earlier. For all domains except processing speed, the FDRs showed deficits when compared to the PHCs. Interestingly, the deficits in social cognition domain for FDRs were comparable with those of MPI group (in Supplementary Material Table 3). Figure 3 shows the estimated marginal means ± 95% confidence interval for the neurocognitive domains among PHC, FDR and MPI.

**Figure 3:**
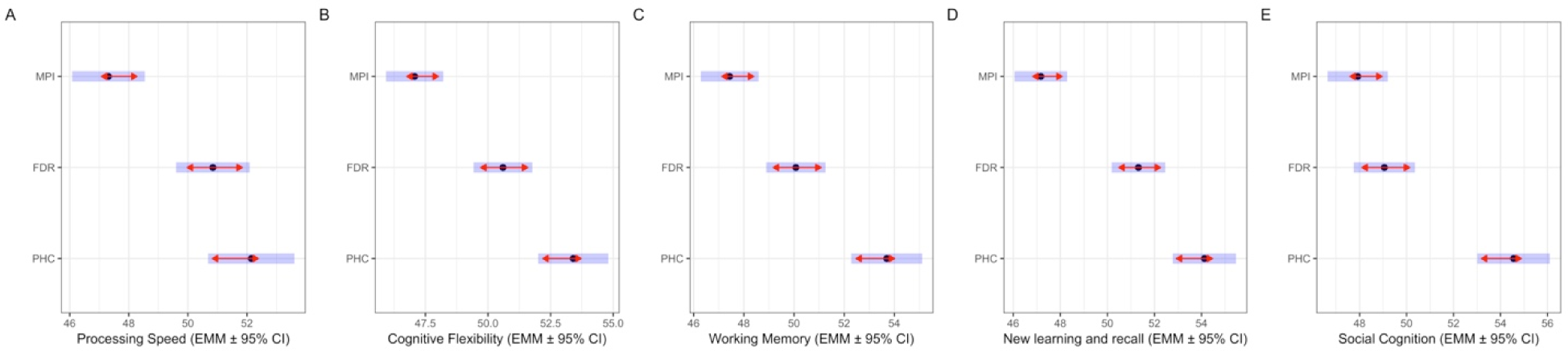
Estimated Marginal Means ± 95% Confidence Interval (CI) for the Neurocognitive domains among Major Psychiatric Illness, First Degree Relatives and Population Healthy Controls. (Processing speed-Panel A, Cognitive Flexibility-Panel B, Working Memory-Panel C, New Learning and Recall-Panel D and Social cognition- Panel E) PHC, population healthy control; FDR, first degree relative; MPI, major psychiatric illness; EMM, estimated marginal means; CI, confidence interval.

### Relationship between neurocognitive components and Family History Density

The diagnosis-specific FHD showed unique relationships with specific cognitive domains. While FHD of SCZ and SUD was associated with significantly poorer scores across all neurocognitive domains, FHD of OCD was associated with significantly poorer cognitive flexibility alone. Individuals with FHD of BPAD had poorer working memory, and more deficits in the verbal memory and recall domains. FHD of Dementia was associated with significantly poorer social cognition (in Supplementary Material Table 4). Figure 4 shows the standardized beta ± standard error for the neurocognitive domains with respective FHDs.

**Figure 4:**
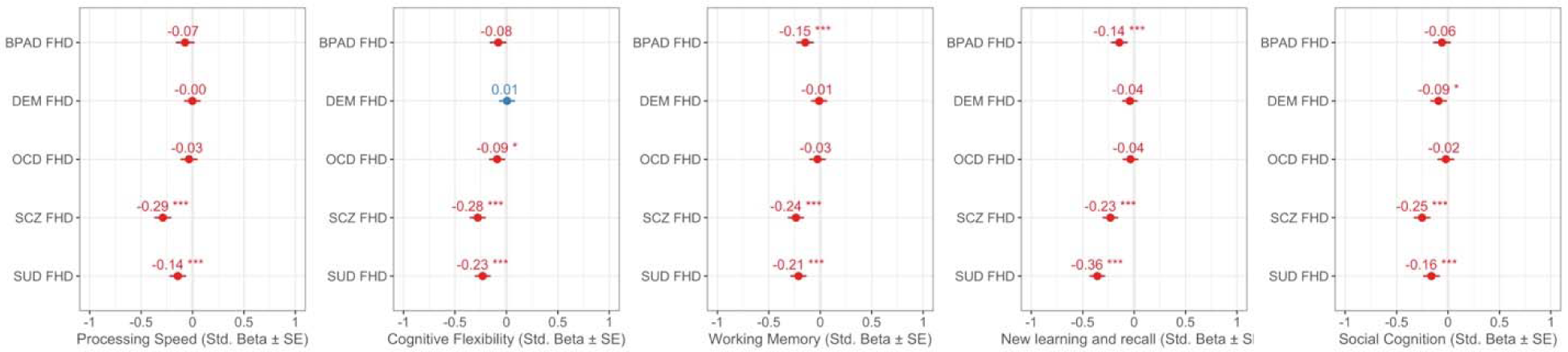
Effects plot representing relationship between neurocognitive components and Family History Density scores. A)Processing speed. B)Cognitive Flexibility. C)Working Memory. D)Verbal(New) Learning and Recall. E)Social cognition FHD, family history density; SCZ, schizophrenia; BPAD, bipolar affective disorder; OCD, obsessive compulsive disorder; SUD, substance use disorder; SE, standard error.

### Relationship between neurocognitive components and illness duration and severity

Within the MPI group, all neurocognitive domains, except social cognition, showed significant worsening with normalized lifetime illness duration (Figure 5), but not with cross-sectional clinical global impression of severity.

**Figure 5:**
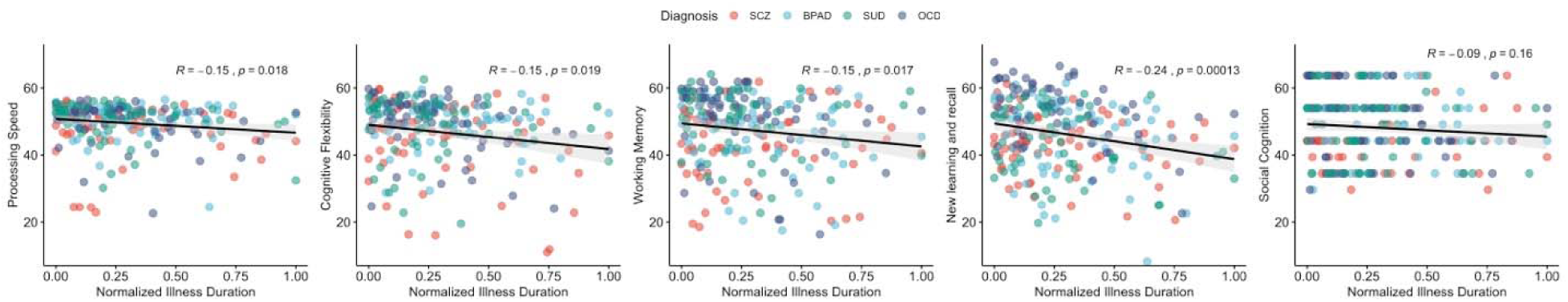
Relationship between cognitive components and normalised illness duration. A)Processing speed. B)Cognitive Flexibility. C)Working Memory. D)Verbal(New) Learning and Recall. E)Social cognition SCZ, schizophrenia; BPAD, bipolar affective disorder; OCD, obsessive compulsive disorder; SUD, substance use disorder

## DISCUSSION

In this study, we aimed to characterize potential transdiagnostic neurocognitive deficits and their endophenotypic value in a large cohort of multiplex families affected with two or more individuals diagnosed with SCZ, BPAD, SUD or OCD.

Consistent with our first hypothesis, we identified transdiagnostic deficits in cognitive flexibility, working memory and social cognition. Verbal memory was affected in SCZ, BPAD and SUD, while processing speed was impaired in SCZ and BPAD. A review of metanalyses of the transdiagnostic neurocognitive deficits revealed that, executive function and episodic memory deficits are transdiagnostic and stable across the life span [47] however we found that these deficits worsened with age. Cognitive control, which mainly involves working memory and cognitive flexibility has been established to be impaired in mental illness across diagnosis with parallel anomalies of brain structure and function [48].This is also broadly consistent with previous meta-analyses from diagnosis specific studies in SCZ and BPAD [49,50],where verbal memory, processing speed and working memory were found to be impaired; SUD [51], where response inhibition, working memory and distractor inhibition control were impaired; OCD [52], where executive dysfunction and non-verbal memory were impaired. Studies where SCZ-BPAD were explored together also reported multiple cognitive deficits across domains[49,53,54]. Our findings support the notion that both diagnosis-specific and transdiagnostic markers are critical in conceptualization of cognitive dysfunction in psychiatric illness [55]. While the diagnosis-specific markers increase the vulnerability to the down-stream effects of specific cognitive dysfunction, the transdiagnostic markers increase an overall risk for general psychopathology that are common across psychiatric illness [56].

As per our second hypothesis, the unaffected FDRs had deficits in all the domains except processing speed in comparison to PHC. The social cognition deficits were comparable to that of affected individuals scores. These deficits thus fulfilled the criteria for an endophenotype indicating that they represent pre-existing familial vulnerability to psychiatric illness. However, in this analysis, FDRs of all the diagnosis were pooled. To see whether these endophenotypes have diagnostic specific validity or whether they are transdiagnostic endophenotypes, we conducted correlation with FHD scores. Division of FDRs across diagnosis groups was impossible because several FDRs had family history of more than one diagnoses (Figure 1). Deficits in cognitive flexibility were significantly associated with FHD scores of SCZ, OCD, SUD; verbal memory and working memory were significantly associated with FHD scores of SCZ, BPAD, SUD; social cognition was significantly associated with FHD scores of SCZ, DEM, SUD. Thus, our transdiagnostic cognitive deficits (i.e., cognitive flexibility, working memory and social cognition) were found to be endophenotypes across disorders.

Similar to our findings, study on children revealed that working memory deficits is a common cognitive liability for mental health disorders [57], particularly externalizing symptoms [58]. Error related negativity which may stem from cognitive inflexibility [59] has been found to be transdiagnostic marker between OCD, anxiety and SUD[60]. Theory of mind, which was particularly evaluated in our study, has neuroanatomical underpinnings of widespread neural networks and has been implicated to be impaired in transdiagnostic neurodegenerative disorders [61]. A systematic review of metanalyses of social cognitive dysfunction across 30 clinical conditions highlighted that the social cognitive deficits are core cognitive phenotypes across psychiatric and neurological illnesses[62]. Comparable social cognition deficits in FDRs to affected individuals could represent apparent difficulty experienced by vulnerable individuals in understanding intentions and emotions, when presented with day-to-day challenges with tasks that demand access to social and conceptual knowledge.

As rightly pointed out by Beauchaine and Constantino, psychiatric disorders are single disease processes that cut across diagnostic boundaries, and need endophenotypic evaluation across disorders based on genetically influenced neuroanatomical basis than specific disorder approach [5]. Our study holds significance in this regard. The only other study which evaluated neuroanatomical basis of endophenotype, concluded that increased putamen volume has transdiagnostic endophenotypic value [8]. However, this had only 20 FDR as opposed to 227 FDRs in our study.

Deficits in these cognitive components were state-independent and did not vary as a function of cross-sectional illness severity (CGI-S) as we hypothesized, but with illness chronicity (life-time duration of illness). We have recruited patients at different stages of illness and majority in their symptom free period which might explain the absence of association between severity of illness and the neurocognitive deficits. The stronger association of deficits with total duration of illness is confounded by uncontrolled variables like duration of untreated illness, effect of treatments. Further controlled studies are needed to establish this relationship.

Transdiagnostic studies when involving multimodal and multivariate approaches[5,55,56,63,64] have the potential to uncover greater signals for psychiatric vulnerability risks, especially for shared phenomena and symptomatology across separate diagnostic entities. These approaches could also aid the discovery of common genetic variation [65] that is relevant to transdiagnostic symptomatology in a manner that is unbiased by traditional nosology. Furthermore, family-based endophenotypic approach for next-generation sequencing employed in the ADBS study [66], could have potential to identify genomic variations across the entire allele frequency spectrum (rare to common), that may contribute to such cognitive impairments.

There are certain limitations that needs to considered. We did not perform a comprehensive neurocognitive battery for all lobe functions but a selected panel of tests due to time constraints. A second limitation is that although we accounted for the effect of demographic variables such as age, parental education and gender in the analyses, we could not account for varying stages of illness and medication status because of difficulty in meaningfully standardizing stages of illness and medications across several psychiatric diagnoses. Nevertheless, this limitation would not affect the endophenotypic analysis, where the unaffected group were unmedicated and did not have clinically diagnosable psychiatric illness. Lastly, the data presented represent a cross-sectional profile of the cognitive dysfunction; the future analyses from the consortium will attempt to model the longitudinal trajectories of cognitive dysfunction along with multimodal brain imaging investigations to understand the neurobiological underpinnings of these dysfunction.

## Data Availability

Data will be made available on reasonable request

## Acknowledgements

The authors are grateful to all the patients, their family members and healthy volunteers who participated in the study.

## Author Contribution

BH contributed to the design of the study, data cleaning and analysing and interpretation, preparation of the initial manuscript; PD, AD, MB, NVM contributed acquisition of the data, analysis and interpretation; DI contributed to data acquisition and revising the work critically for important intellectual content; SB, JM, VSS contributed to design of the study, data acquisition and interpretation of the results; VB, JYCR, UMM contributed to design of the study; ADBS Consortium contributed to design of the study and data acquisition and approved the final manuscript; BV contributed to design of the study, moderating the data acquisition, planning of the analysis, interpretation of the results and manuscript preparation and revision and final approval.

